# Possible harms of *Momordica charantia L*. in humans; a systematic review

**DOI:** 10.1101/2022.10.22.22281390

**Authors:** Armelle Demmers, Jurriaan J. Mes, Roy G. Elbers, Raymond HH Pieters

## Abstract

**Introduction:** A few cases of serious side effects have been reported of Momordica charantia L. (MC). No comprehensive safety assessment has yet been performed based on human intake.This systematic review aims to evaluate the potential harm of Momordica charantia L. derived products using data from randomized controlled trials.

**Methods:** Databases Cochrane Library, Pubmed and EMBASE were searched until December 2020. The PRISMA harms checklist was followed. Data extraction was on aspartate aminotransferase (AST), alanine aminotransferase (ALT), creatinine, adverse effects (AE), reasons for drop out related to the intervention and interaction with other treatment. Two authors independently extracted data and bias was evaluated based on the latest version of the Cochrane risk of Bias Tool (RoB 2). Additional safety data were requested from Health Regulatory Agencies, Herbal Medicine Associations and manufacturers.

**Results:** Seventeen trials met the inclusion criteria. The IRR was calculated for each study ranging from 0.30 (95% CI = 0.12 to 0.75) to 13.00 (95% CI = 0.73 to 230.76) of any adverse events.

**Conclusions:** Under a daily dosage of 6g of MC-derived products no evidence was seen of harms in humans. In case reports that showed serious harm, MC was used in a liquid form. The safety of traditional MC-based supplements appears more guaranteed when produced under strict quality standards.

## 1 Introduction

Traditionally Momordica charantia L. (MC, bitter lemon) is used for eczema, psoriasis, cancer, rheumatism, antiviral activities and to control blood glucose levels (Grover & Yadav, 2004; Mahomoodally & Ramalingum, 2015; Panigrahi et al., 2010). These indications have been supported with animal and in vitro studies (Bai et al., 2016; Bai et al., 2018; Bhat et al., 2018; Huang et al., 2013; Joseph & Jini, 2013; Kim & Kim, 2011). Recently, two systematic reviews were conducted to evaluate the effect of MC on control of blood glucose levels in human and these reviews showed conflicting results (Ooi et al., 2012; Peter et al., 2019). No statistically significant difference was found in the Cochrane systematic review with regard to the glycaemic control when effects of MC preparations were compared to placebo (Ooi et al., 2012). On the contrary, in the other systematic review MC formulation was found to significantly reduce fasting plasma glucose (FPG) levels, postprandial glucose (PPG) and haemoglobin A1c (HbA1c) when compared to placebo (Peter et al., 2019). The methods of these systematic reviews were different with regard to inclusion of polyherbal supplements, randomization and minimal follow-up time.

Several case reports on safety issues have been published (Hullin, 1988; Erden et al., 2010; Nadkarni et al., 2010; Mohanty et al., 2018) which justifies a closer analysis of safety.

MC seeds contain vicine-like compounds which may induce a hemolytic disorder known as favism (Raman & Lau, 2011). Heart and blood disorders have been observed in animals as well (Khan et al., 2019, Abdillah et al., 2019, Temitope, 2014).

Significant increases of liver enzymes have been observed in rats treated with fruit juice and seed extract in different intervention groups (Tennekoon et al., 1994). Furthermore, extensive inflammation and cell toxicity occurred in the white adipose tissue of mice subjected to high doses of bitter melon seed oils (Chen et al., 2012).

Further, reproduction problems have been observed in animal studies (Adewale et al., 2014, Odewusi et al., 2010). Also, in dogs fed with total fruit extract (1.75g/ day oral for 60 days) anti-fertility and anti-spermagenic effects have been found (Dixit et al., 1978). Various parts of the plant have been shown to cause malformations of embryos in pregnant animals (Uche-Nwachi & McEwen, 2010; Yeung et al., 2009, Khan et al., 2019). It is not yet clear which components may cause these effects, but it seems that specifically two enzymes, β-and α-momorcharins, may be abortifacient (Yeung et al., 2009). The question arises if these animal findings can be extrapolated to humans and which plant part could cause these effects. In addition to the possible bioactive substances of the plant, the exact plant material of the product may also play an important role. Most of the adverse events (AE) of herbal products and herbal medicines can be attributed to poor quality, processing and (im)purity of the product (Ahmed et al., 2019; Chan, 2003). This stresses the need for a critical systematic review of the available data on safety of MC use in humans. Here, we evaluate the AE reported after MC use, in relation to the plant material and the daily dosages of the MC-derived products.

### 1.1 Objective

A systematic review of available data to determine harms of the use of Momordica charantia L. derived products in humans.

## 2 Methods

The PRISMA harms checklist was followed (Zorzela et al., 2016). When side effects or AE were described as outcome measures in the protocol or the phrase ‘there were no AE’ was reported, the trial was included. When the study report did not mention anything side effects or AE, the study was excluded.

Additional safety data were obtained from international drug monitoring agencies, manufacturers and distributors of MC, and herbalist organizations.

### 2.1 Selection criteria

Randomized controlled trials (RCTs) with any kind of intervention period were included but only those published in English. We included RCTs with healthy participants and with participants displaying any kind of disorder. RCTs mentioning any oral therapy using single MC in any dosage or formulation were included. Studies using a combination with MC and other herbal(s) or food ingredients preparations were excluded. Interventions in the control group could be: no treatment, placebo, or any other treatment.

### 2.2 Search methods

We searched the following electronic databases until December 2020: The Cochrane Library, Pubmed and EMBASE. The search plant names were “Momordica charantia” or “bitter melon” or “bitter gourd” or “bittergourd” or “balsam pear” or “bitter squash” or “karela” or “amplaya” or “sopropo” in subject, abstract and keywords. Unpublished or ongoing trials were searched in the electronic databases until December 2020 clinicaltrials.gov and the World Health Organization International Clinical Trials Register Platform.

Authors of unpublished trials were asked for data by email. Hand search in Google Scholar, Mendeley, ResearchGate and reference lists of reviews was performed by A.D.

### 2.3 Data extraction

The results from the searches were independently screened by author A.D. and R.P. Titles and abstracts were scanned on inclusion and exclusion criteria. Full text investigation was performed when titles and abstracts gave insufficient information. In case of disagreement a third author (J.M.) was consulted to reach a final decision.

From the selected studies the following data were extracted:

(a) General study information: authors, publication year, samples sizes, follow-up period, methods of AE assessment; (b) Intervention data: plant material, administered dosages, administration frequency and duration, intervention control group; (c) Results on safety parameters: AST, ALT, creatinine, all reported AE, reasons for drop-out related to the intervention, interaction with other treatment, information about time between intake and adverse-event, number of participants who experienced an AE, follow-up time/time being at risk; (d) Baseline characteristics of subjects: age, sex, body mass index (BMI), diagnose and use of medication.

### 2.4 Assessment risk of bias

The validity of the study results was assessed by A.D. and J.M. using the latest version of the Cochrane Risk of Bias Tool, a checklist evaluating the validity of studies as to the following five bias parameters; the randomization process, deviations from intended interventions, missing outcome data, measurement of the outcome and selection of the reported result (Sterne et al. 2019).

We assessed bias of starting and adhering to the intervention instead of intention-to-treat because of AE as outcome. For the judgements of the parameter of outcome measurement we rated the methods of AE assessments used in the trials.

### 2.5 Statistical analyses

We conducted meta-analyses in R-studio with the package Meta (Balduzzi et al. 2019). Where statistical pooling was not sensible to clinical heterogeneity, the effect size of each study was separately presented in a forest plot. For continuous data, differences between the control and intervention groups were calculated based on mean difference. AE were analysed as dichotomous data. For comparisons of AE incidence rate ratio (IRR) was calculated with the corresponding 95% confidence intervals (95% CI) using data extracted. The IRR was calculated as follows: dividing the number of AE by the number included participants in the study x time of study period in days.

All reported AE were analysed as total numbers of occurrence because multiple episodes of a side effect could have occurred in one participant. For each reported AE a separate analysis was performed. Similar and most common AE were clustered.

Drop-outs were analysed separately and also counted as an AE if the report was clear or if the assumption that the drop-out was related to the intervention was plausible. When a study reported no events, 0.5 was added to all four cells of the 2×2 table.

When multiple intervention groups from one study were in a meta-analysis, we combined the groups to create a single pair-wise comparison. We established a cut-off value of 3g per day of MC to analyse as a separate group because most AE were seen in trials with a daily dose of > 3g MC per day (Cortez-Navarrete et al.,2018; Rosyid et al., 2018; Soo May et al., 2018).

## 3 Additional safety data

Additional safety data were obtained from Healthcare Regulatory Agencies, manufacturers and distributors of MC, and herbalist organizations worldwide. Also, we searched for case reports on safety in the databases Pubmed and EMBASE for available literature and to traditional knowledge from ancient use about safety of MC.

## 4 Results

### 4.1 Results of the search

The database searches and additional records gave a total number of 239 studies. After removing duplicates, 85 studies remained. We also included an unpublished trial conducted by author J.M. of this review as it met the inclusion criteria (Mes et al., 2020). The flow diagram (Fig. 1) shows that 56 studies did not met the inclusion criteria by reading the title or the abstract and that we retrieved the full-text papers of 30 studies for further assessment. In five of these 30 remaining studies, no safety issues or AE were reported (Boone et al., 2017; Kasbia et al., 2009; Kinoshita et al., 2018; Inayat-ur-Rahman et al., 2019; Selvakumar et al., 2017). The full-text of six of the 30 studies could not be obtained (Amirthaveni et al., 2018; Kim et al., 2018; Muthumani & Ahmed John, 2009; Pongnikorn et al., 2003; Shi et al., 2004; Tayyab et al., 2013) and we were unsuccessful in our efforts to obtain these articles directly from the authors. We were also unsuccessful in getting information from the authors on reports that did not report any safety parameters or AE.

**Figure 1.**
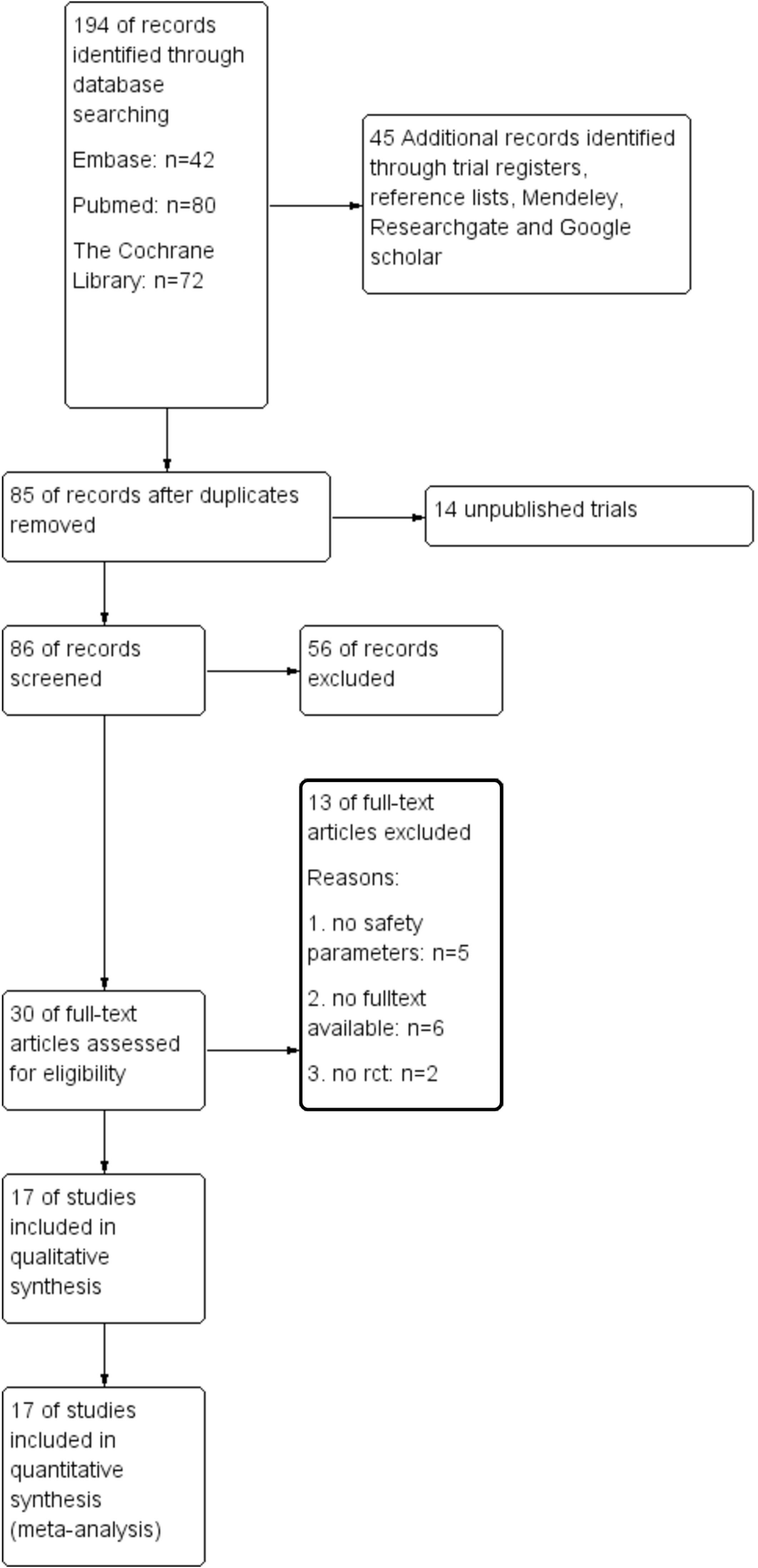
Flow diagram

Finally, seventeen RCTs were included for analysis (Table 1). In our analyses of one trial (Zänker et al., 2012) we excluded the intervention group with 100μg chromium and 10mg zinc as intervention next to MC because of the inclusion criteria.

**Table 1.** Baseline characteristics

| References | Sex<br>(male:female) | Age in<br>years<br>(mean) | Body Mass<br>Index | Used medication<br>next to MC | Diagnose of the<br>participants |
| --- | --- | --- | --- | --- | --- |
| Cortez-Navarrete 2018 | 33:67% | 48.5 | 28.9 | none | Type-2 diabetes |
| Dans 2007 | unclear | 59.2 | 26.2 | 100% anti<br>diabetics | Type- 2 diabetes |
| Fuangchan 2010 | 26:74% | 51.8 | 25.1 | none | Type- 2 diabetes |
| Hsu 2020 | unclear | 62.9 | unclear | 100% anti<br>diabetics | Type- 2 diabetes |
| John 2003 | 32:68% | 53.2 | unclear | 100% anti<br>diabetics | Type- 2 diabetes |
| Kim 2020 | 38:24% | 58.1 | 25.3 | 100% anti<br>diabetics | Type- 2 diabetes |
| Krawinkel 2018 | 46:54% | 47.8 | 29.6 | none | prediabetes |
| Kumari 2018 | unclear | unclear | 27.7 | 100% anti<br>diabetics | Type-2 diabetes |
| Lim 2010 | 45:55% | 56.6 | 25.7 | none | Type- 2 diabetes |
| Mes 2020 | 70:30% | 67.8 | 28.7 | none | prediabetes |
| Rahman 2015 | 66:34% | 52 | 25.45 | none | Type- 2 diabetes |
| Rosyid 2018 | 33:67% | 53.7 | 21.6 | 100% anti<br>diabetics,<br>100% anti<br>biotics | Type-2 diabetes<br>with diabetic foot<br>ulcer |
| Soo May 2018 | 30:70% | 59.9 | 26.9 | Analgesia allowed | primary knee<br>osteoarthritis |
| Suthar 2016a | 62:38% | 41.3 | unclear | 89% anti diabetics | Type- 2 diabetes |
| Suthar 2016b | 56:44% | 48.55 | 27.29 | none | Type- 2 diabetes |
| Trakoon-osot 2013 | 29:71% | 57.9 | 25.7 | 76% anti diabetics | Type- 2 diabetes |
| Zanker 2012 | 72:28% | 62.6 | 29.3 | 77% anti diabetics | Type- 2 diabetes |

### 4.2 Characteristics of included studies

The 17 studies included a total of 1.320 participants, varying from 24 to 142 in number per study and with a total 785 participants in the MC intervention group and 535 participants in the control group. The participants had a mean age between 41.3 and 67.8 years. Baseline characteristics concerning age, sex and BMI of subjects are reported in Table 1.

The mean intervention period was 67 days, ranging from a single day (1 dose) to an 112 days intervention period. Four studies did not report details about the production process of the MC derived product (Cortez-Navarrete et al., 2018; Dans et al., 2007; John et al. 2003; Lim et al., 2010).

The plant material used in the various studies was very different. Some studies investigated the supposed active constituent charantin which is associated with blood sugar lowering properties (Fuangchan et al., 2011; Rahman et al., 2015; Trakoon-osot et al., 2013; Zänker et al., 2012). In Rahman et al. the amount of charantin per daily dosage was not clear. For two studies the extraction of MC juice was dried and powdered (Suthar_a et al., 2016, Suthar_b et al., 2016). No information has been reported about the used plant material in four studies (Fuangchan et al., 2011, Hsu et al., 2020, Soo May et al., 2016., Kumari et al.,2018). Other details on the included studies can be found in Table 2.

**Table 2.**
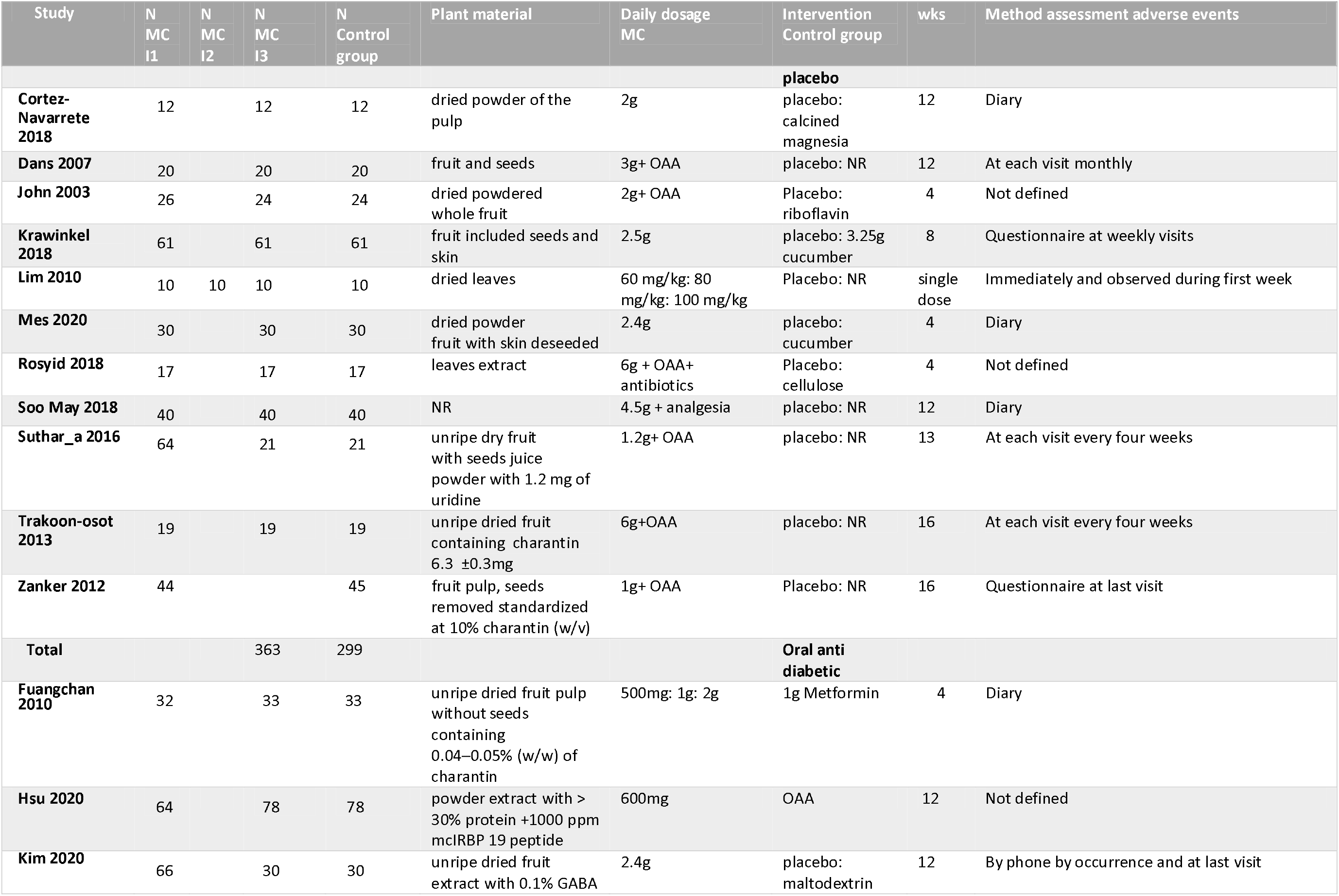
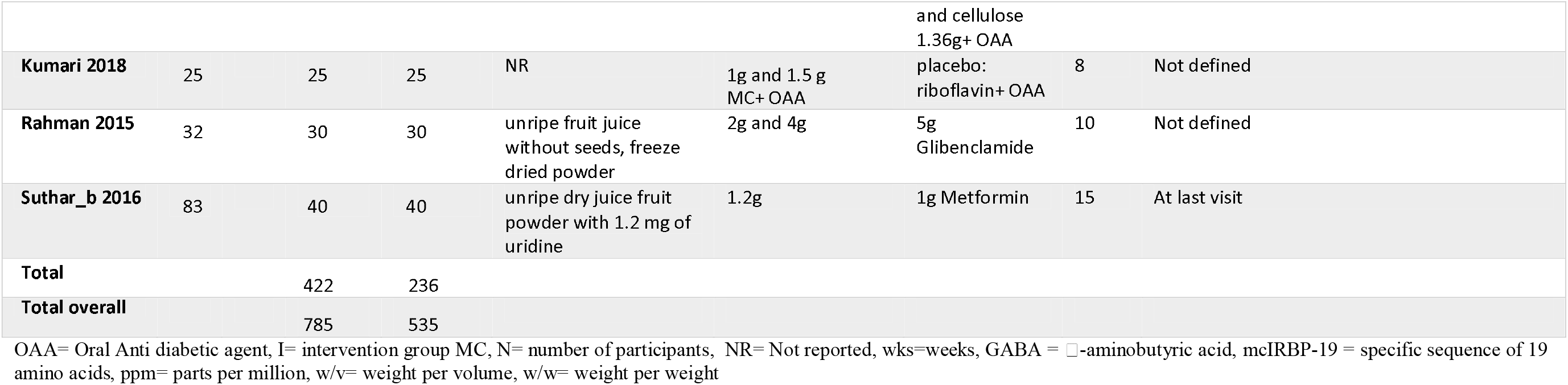
Description of included studies

### 4.3 Risk of bias

Risk of bias was assessed for the outcome AE (Fig 2). In one study (Hsu et al., 2020) AE were measured by liver and kidney enzymes. We analysed these indicators as separate outcomes, not as AE in general. Therefore, we excluded this trial in the risk of bias assessment.

**Figure 2.**
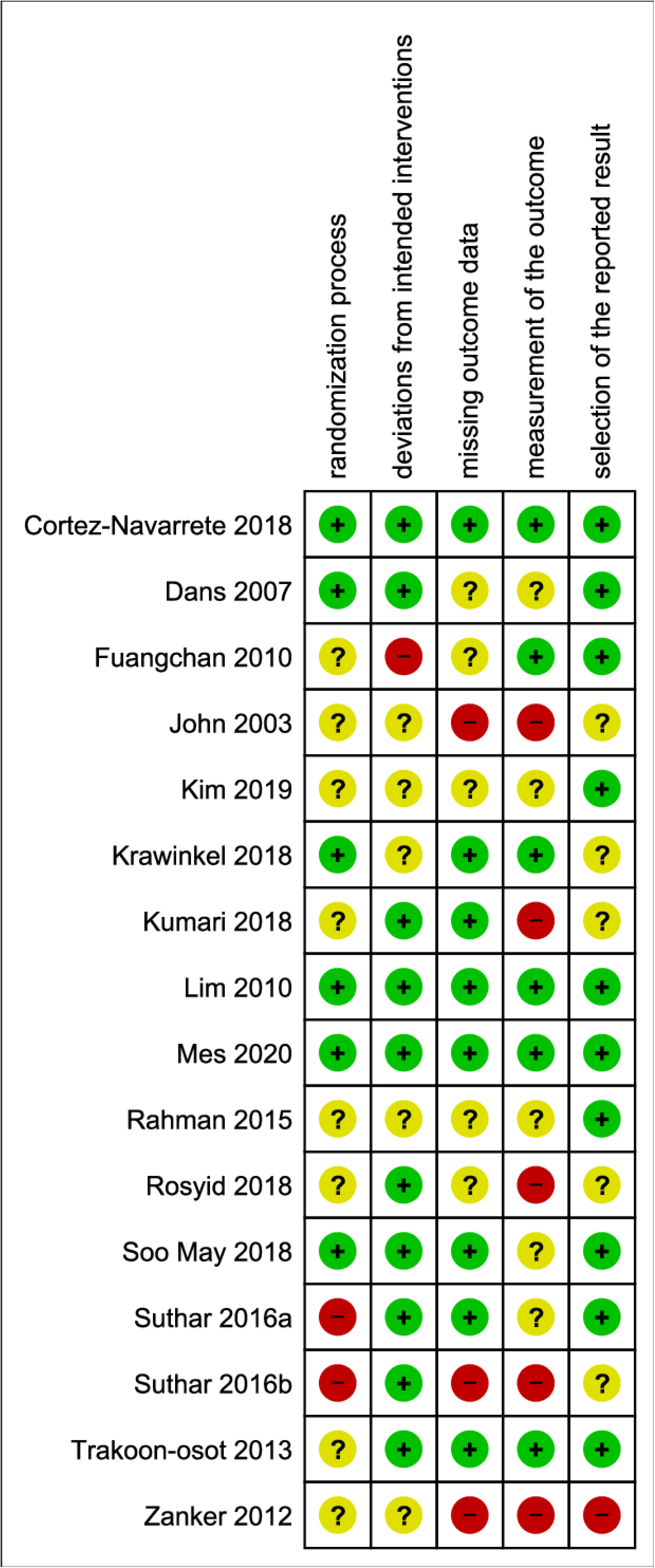
Risk of bias summary

For the domain “outcome measurement”, nine studies were unclear with regard to AE. These were consistently tracked or only reported occasionally (Dans et al., 2007; John et al., 2003; Kim et al., 2020; Kumari et al., 2018; Lim et al., 2010; Rahman et al., 2015; Rosyid et al., 2018; Suthar_b et al., 2016; Zänker et al., 2012).

In one trial, the reporting of AE for the domain “selection of the reported result” was no reporting at all if there were any AE reported or not (Zänker et al., 2012). We judged this trial as highly biased.

### 4.4 Analysis of the comparisons

Due the strong clinical heterogeneity between the studies with regard to characteristics of participant, plant material, and dosage, it was not possible to conduct meta-analyses. We decided to graphically present the results in forest plots, but we refrained from estimating pooled summary statistics.

Only five RCTs reported a comparison with placebo and these are used in the forest plot of figure 3.

**Figure 3.**
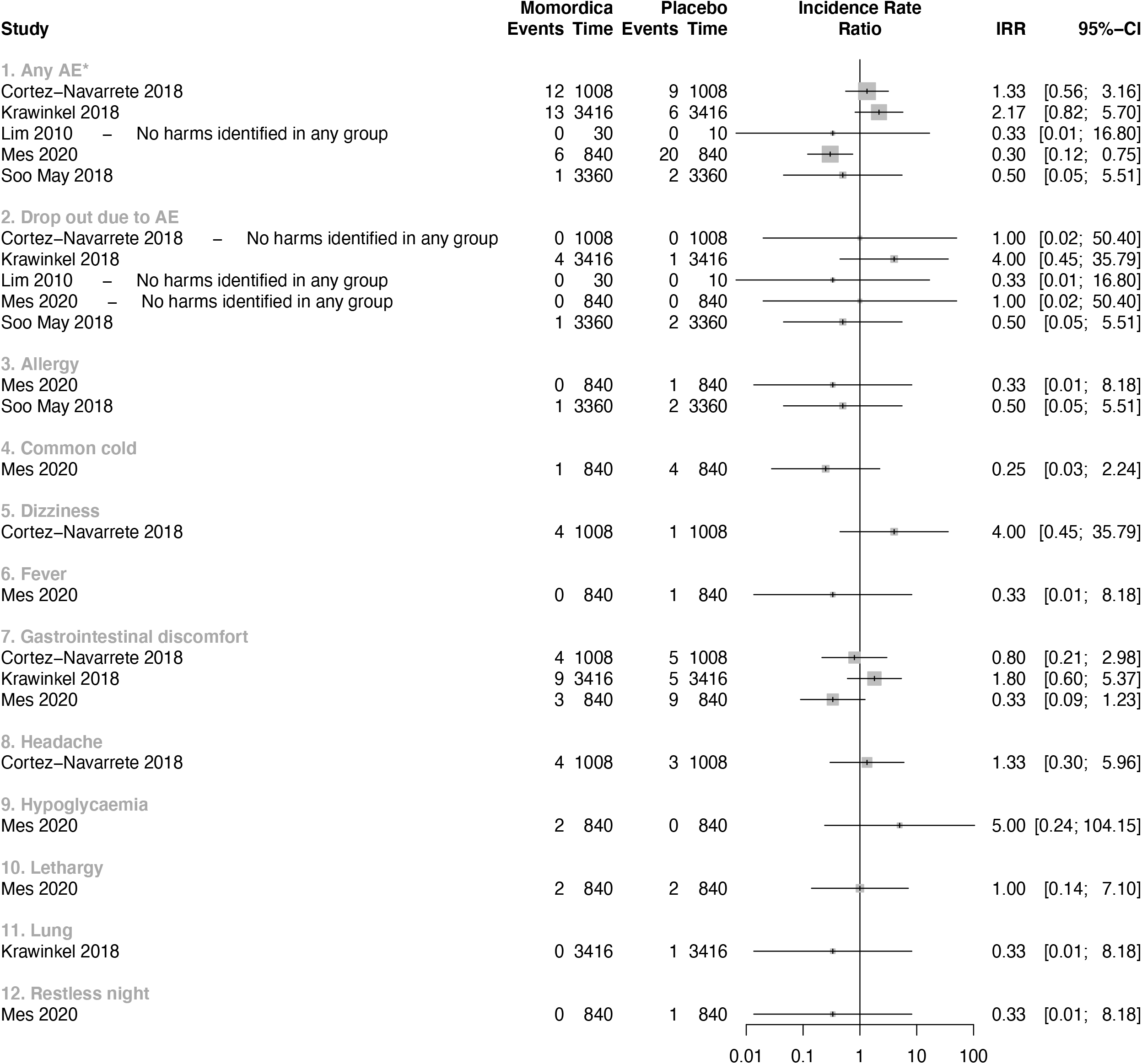
Forest plot MC compared with placebo

Figure 4 summarizes the results of three studies that compared MC with usual treatment. One study (Rahman et al., 2015) had two intervention arms, one with 2g MC daily (a) and one of 4g MC daily for ten weeks (b). We analysed these two arms separately. In a study that included T2DM patients (Fuangchan et al., 2011), increased appetite as AE was found after intake of 2g/day MC more frequently than in the groups with 500mg/day and 1g/day compared to the Metformin treatment groups.

**Figure 4.**
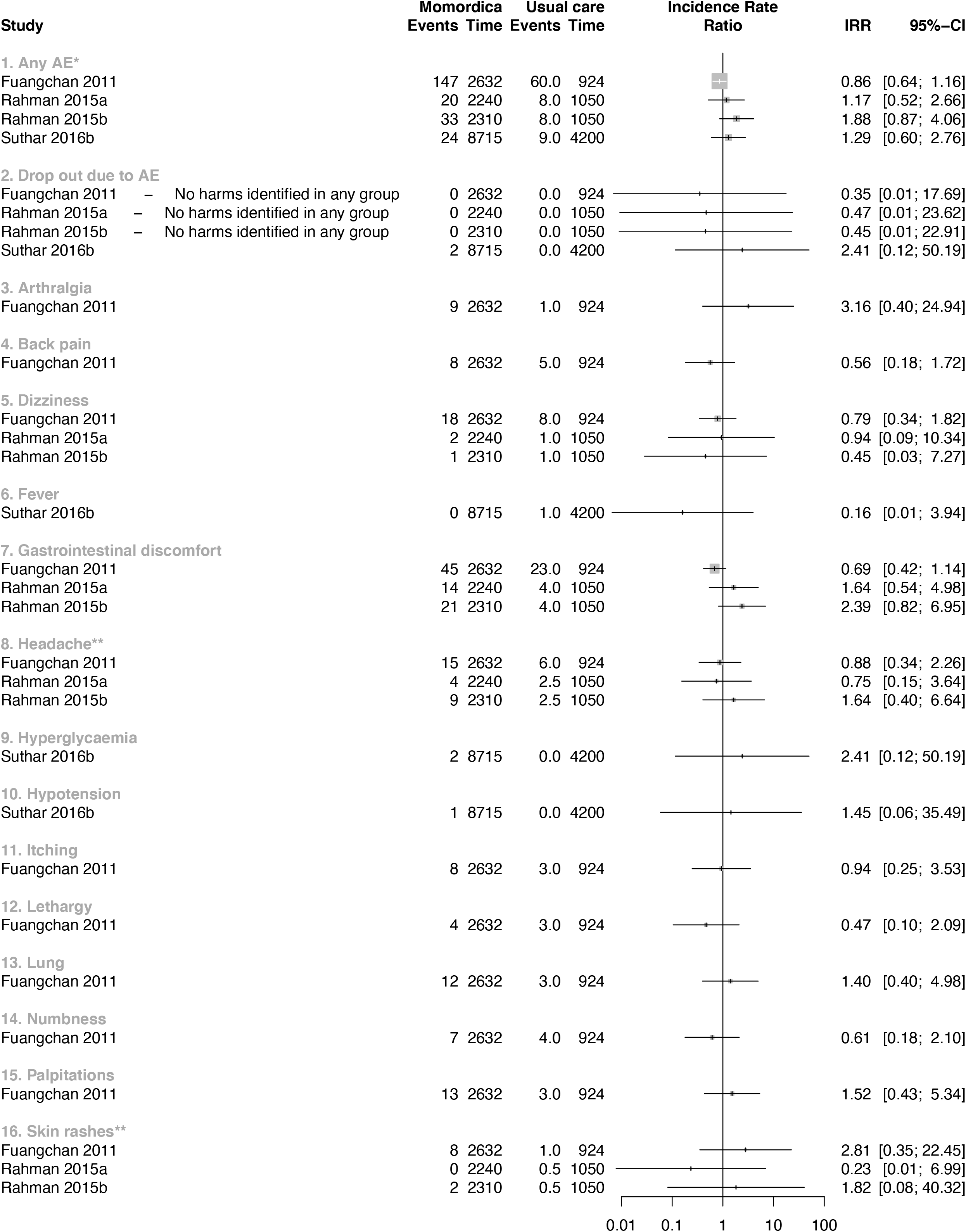
Forest plot MC compared with usual treatment

Other AE were 16 episodes of palpitation. Few of these episodes were associated with hypoglycaemia. These symptoms resolved with rest and did not require any treatment or discontinuation of Metformin or MC. In another trial T2DM participants of 4g/ day MC group had more appetite and headaches compared with 2g/day or Glibenclamide 5mg/day (Suthar_b et al., 2016).

Seven studies were included in the comparison between MC as co-intervention compared with placebo (Fig 5). All seven studies administered anti-diabetic medication as co-intervention. In one of these studies participants used also antibiotics (Rosyid et al., 2018). The IRR was calculated for each study, ranging from 0.33 (95% CI = 0.01 to 16.54) to 13 (95% CI = 0.73 to 230.76) for AE.

**Figure 5.**
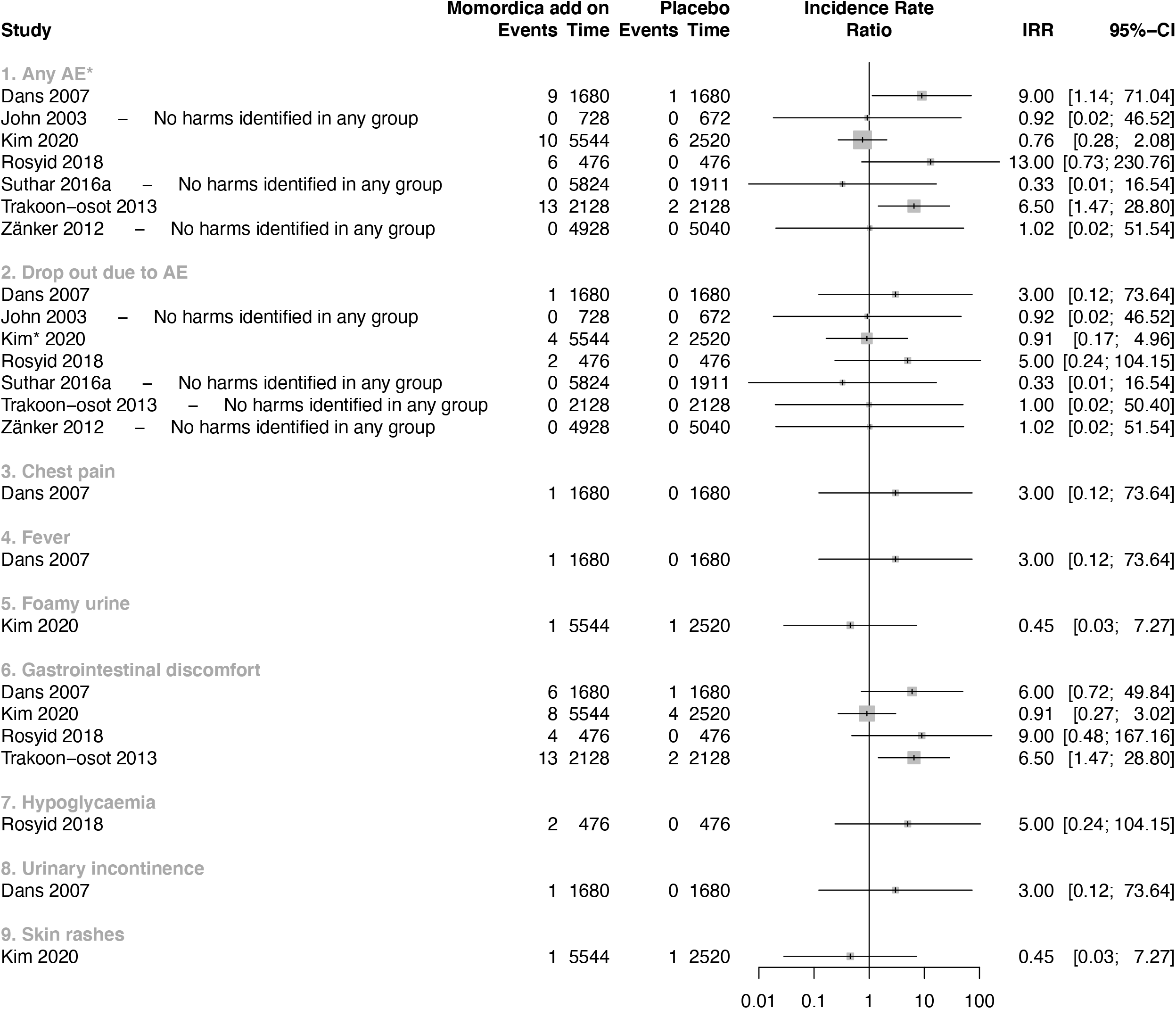
Forest plot MC as additional co-intervention compared with placebo

Two studies showed a large effect on the overall AE outcome. The wide CI’s showed the uncertainty of these IRR’s values. The used plant material in one of the study was prepared from unripe dried fruit without seeds, and administered in a high daily dosage of 6g. At this dosage, more gastrointestinal discomfort with symptoms of diarrhoea and flatulence with an IRR of 6.50 (95% CI = 1.47 to 28.80) was found (Trakoon-osot et al., 2013). In the other study, a daily dosage of 3g fruit with seeds was used (Dans et al., 2007). In a study that included people with T2DM and diabetic foot ulcer, two participants dropped -out because of nausea, vomiting and hypoglycaemia. In this study a dose of 6g leaves extract of the MC was used in combination with standard medication (Rosyid et al., 2018).

In one RCT MC was studied as additional co-intervention next to Metformin and Glibenclamide and compared with standard oral anti-diabetic agents and placebo (Kumari et al., 2018). The authors of this study reported AE but did not mention in which intervention arm the AE occurred. We assumed that these happened in the MC arm and included them in the data analysed as such (Fig 6).

**Figure 6.**
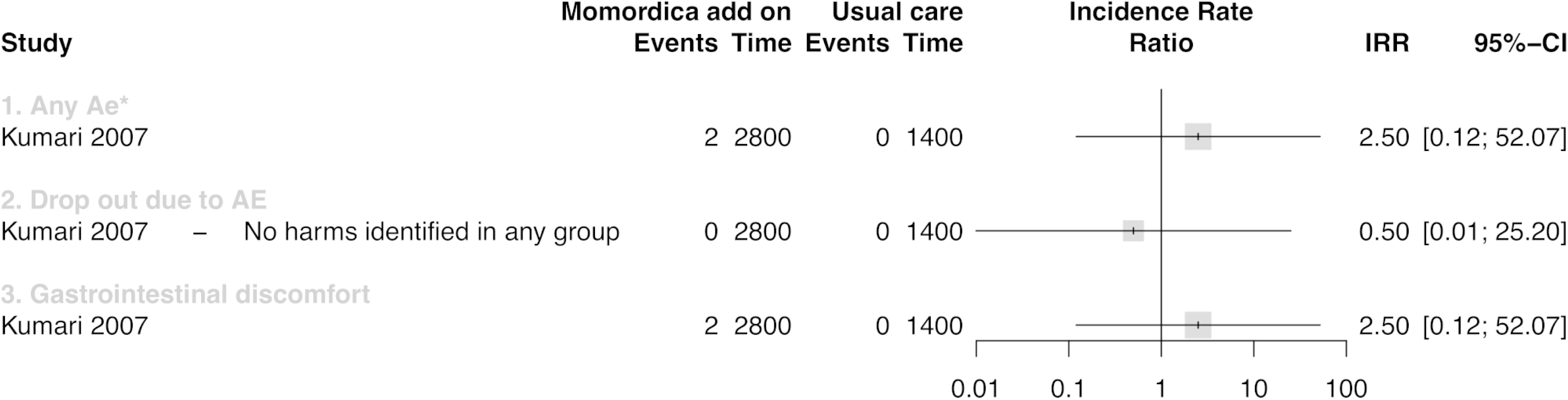
Forest plot MC as additional co-intervention compared with usual treatment

Two notifications of gastrointestinal discomfort were the only AE (Kumari et al., 2018).

### 4.5 Safety parameters

Only one study reported the standard deviations of the means (Dans et al., 2007), for the other studies we imputed these following the methods advised by Cochrane (Follmann et al., 1992). Figure 7 summarizes the results for the safety parameters. Two studies reported that there was no significant influence of MC on renal and liver function, but did not present detailed numbers (Krawinkel et al., 2018; Kim et al., 2020). In one study creatinine was reported in mmol/L instead of μmol/L (Cortez-Navarrete et al., 2018). The author confirmed this. In another study the converted creatinine mean value of the control group (247.60μmol/L) seems much too high (Hsu et al., 2020). We assumed that this reporting was not correct and notified the author. We excluded this study for analysis after a time period with no reaction of this author.

**Figure 7.**
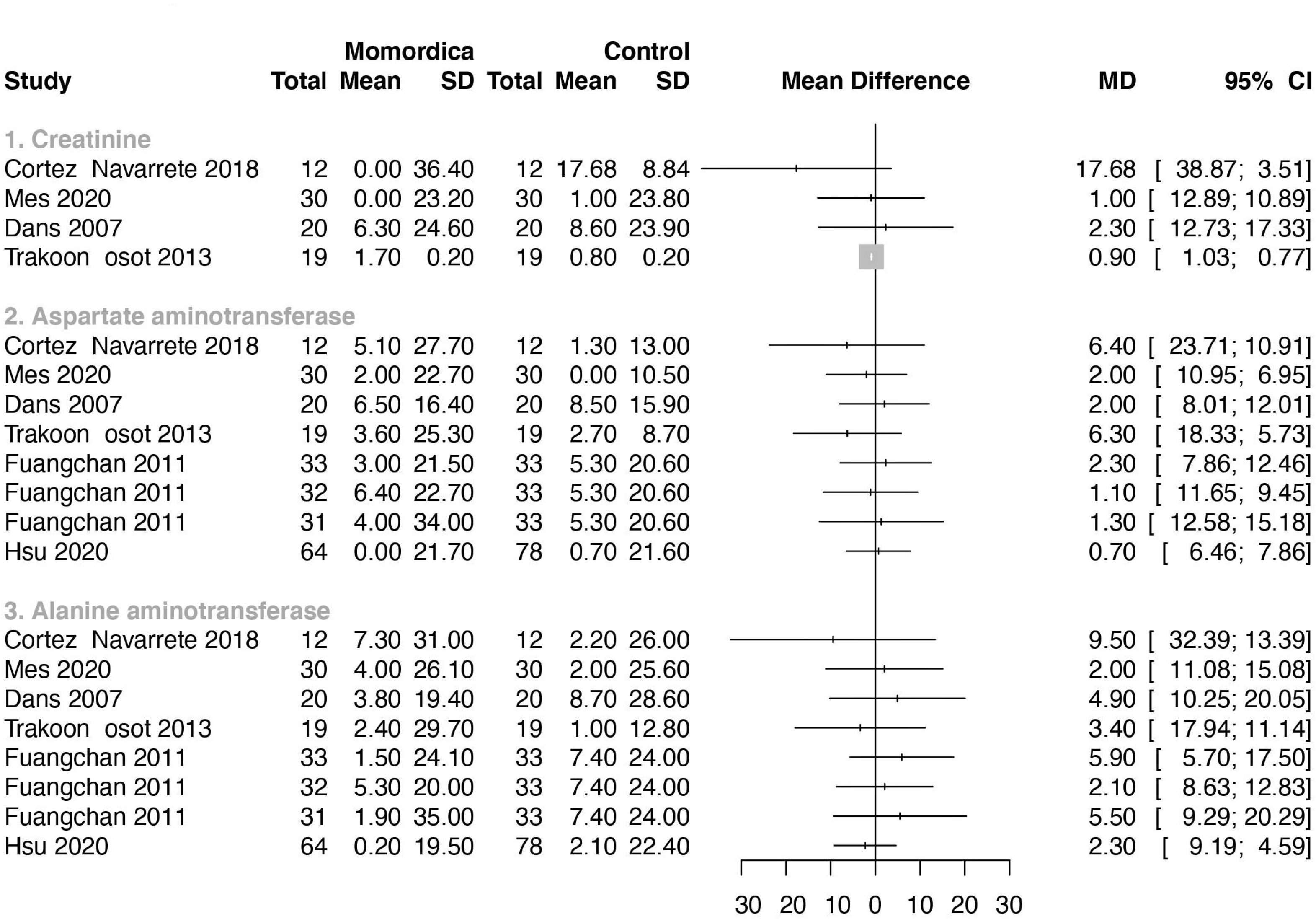
Forest plot ALT, AST, creatinine

ALT and AST were used as outcome parameter in seven studies (Fig 7). In one study no values were reported (Zänker et al., 2012). No statistical significant differences were found in levels of ALT and AST between groups.

### 4.6 Results of additional safety data search

In the FDA center for food safety and applied nutrition (CFSAN) AE reporting system (CAERS) we found three reports of AE after using MC. There was a case of a patient in a life-threatening situation, visiting a healthcare provider with symptoms of vomiting, nausea, malaise, hypersensitivity, haemorrhage, haemoptysis, diarrhoea, asthenia and abdominal upper pain. The suspected cause was a dietary supplement of Nature’s Herbs with bitter melon containing 525mg fruit. The second case report is of a 60-year-old patient that visited a healthcare provider with hypertension. The suspected cause was a rapid release supplement of Puritan’s Pride with bitter melon fruit 900mg daily. During the same period, this subject consumed many other products that could be the cause of the symptoms. The last case was a patient with AF who visited a healthcare provider after the use of Puritan’s Pride bitter melon, 450mg rapid release capsules, twice daily. Detailed information about Puritan’s Pride bitter melon could not be found on the manufacturer’s website.

VigiBase reports refer to a suspected but not confirmed causal relationship between a drug and an event. In the Vigibase of the WHO, eight cases of adverse reactions to MC were found.

In the Pubmed and EMBASE databases we found four case reports. There is a report of a potentially fatal reaction in two young children. They fell into hypoglycaemic coma after drinking tea of MC leaves (Hulin et al., 1988). Another case report is of a 22-year-old man of AF due to drinking MC juice. He consumed crushed MC (Erden et al., 2010). A 42-year-old man admitted to the Emergency Department had complaints of fatigue, intermittent dark urine for 1 week, fever, chills, vomiting and loose black stools for 1 day. He mentioned increased consumption of Chinese MC tea for his hyperlipidemia for 11 days prior to hospitalization. This man was G6PD-deficient and therefore some drugs, foods and chemicals can trigger haemolysis (Mohanty et al., 2018). Another case report involved a 40-year-old man with severe epigastric pain and hematemesis (around 200-300 mL) within half an hour following drinking half a litre of liquid extract of MC (Nadkarni et al., 2010). It is unknown how many supplements of MC are used worldwide a year.

## 5 Discussion

A MC-derived product should be safe for humans and not induce any risk. We performed a systematic review to identify possible harms of MC as supplement based on previously conducted RCTs. Our analysis indicated a strong heterogeneity between the included studies on used plant material, daily dosages, intervention period and participants. Therefore, we created an overview and calculated the IRR separately for each included study in a systematic way but did not combine these results in meta-analyses. Two studies with 6g daily dosage potentially indicate that a too high amount of dried fruit, seeds and leaves might cause a health risk (Rosyid et al., 2018, Trakoon-osot et al., 2013). Some of the AE reported by the subjects, like dizziness, headaches, increased appetite, lethargy, lung problems, palpitations, nausea, constipation and vomiting, could be related to the high and fluctuating blood sugar levels as in many studies people with T2DM were included (Rosyid et al., 2018; Fuangchan et al., 2011; Suthar_b et al., 2016).

A systematic review of RCTs is a good basis for toxicological risk evaluation when enough data can be included and combined in a meta-analysis. The strength of this systematic review is that it was performed with seventeen included RCT’s with the effect size IRR on AE which calculated the harmful effect over time. However, RCTs will not identify any harm on fertility or reproduction problems, which may be a concern since MC has been found to cause reproduction problems and teratogenic effects in animals (Adewale et al., 2014, Dixit et al., 1978, Khan et al., 2019, Odewusi et al., 2010, Yeung et al., 2009). Also, traditional healers in India and Africa have used the seeds of MC to induce abortions (Beloin et al., 2015). Bitter melon seeds contain vicine-like compound which may induce G6PD-deficiency resulting in breakdown of red blood cells (Raman & Lau, 2011), which is indicative of harmful effects of seeds.

We found four case reports of harm in the databases Pubmed and EMBASE. These case reports of AE were not after intake of MC as dietary supplement but after drinking large amounts of juice or tea. This may suggest that liquid use of MC has a different absorption than when taken as a dietary supplement.

The European Food and Safety Authority (EFSA) has concluded: “Safety of an herbal preparation can be presumed when available data would allow concluding that exposure to known levels of the botanical ingredient has occurred in large population groups for many years without reported adverse effects” (EFSA, 2009). The fruits of MC are consumed over decades at different regions in the world without any reported safety concern. So, it can be assumed that when intake does not exceed an equivalent of what can be consumed as vegetable in a meal it would be fair to conclude safety of MC.

Based on our findings concerning the high risk of bias due to missing outcome data and outcome measurement, we would like to stress the importance of systematic collection and detailed reporting of AE during intervention trials. We encountered many incomplete reporting on compliance and reasons for lost to follow-up which can have affected our results. Time to event information could confirm that an reported harm is associated with the intervention. Information about time-to-discontinuation or time-to-withdrawal for each study group was only reported in one trial (Dans et al., 2007). Also, information about severity of the complaints were not reported. If numbers of AE were provided, details about on how many days and in how many people events occurred were missing. A consequence of this limited information was that a causal relationship, in line with the Bradford Hill criteria (Hill, 2015), between the AE and the use of MC could not be established

In five studies selected for this systematic review, the researchers did not provide any information on quality assessment of the product and therefore AE due to contaminants could have influenced our results. Good Manufacturing Practice (GMP) is one of the most important tools to ensure quality of pharmaceuticals and herbal medicines (WHO, 2007). For research but also for safety reasons, GMP should become an important standard for medicinal plants derived products. Plant material can be contaminated with toxic plants containing specific alkaloids (De Wit et al., 2021). Especially microbial safety is of high importance to include in all these studies as food pathogens like E.coli, Salmonella and Listeria can cause effects within hours after intake (Alwakeel 2009). A microbiological and pharmaceutical quality and safety assessment on over-the-counter herbal weight loss supplements in Egypt showed that, based on microbial count, 100% of the unapproved weight loss products had poor bacteriological quality (Ahmed et al., 2019). To date, the European Pharmacopoeia TCM working group has elaborated about 80 TCM herbal monographs with ISO standards for over 14 MPs published or under development. These standards provide important references to the quality consistency and safety of MPs (Xiong et al., 2021).

So, with the limitations of this systematic review, no clear evidence was found that MC preparation and available supplements show more harms than placebo or commonly prescribed OAA under a daily dosage of 6g of dried fruit or leaves. Next to that avoid using high amounts of concentrated MC in juice or tea.

## 6 Conclusion

Comparing collected results and ancient use, MC-based supplements can be assumed to be safe under a daily dosage of 6g. Due to incomplete reporting on compliance and reasons of lost to follow-ups of the participants necessary information is missing to draw a complete conclusion of harms. Causal relationship cannot be confirmed between the AE of the found cases and the intake of MC because of lack of required information. MC is not recommended during birth wish, pregnancy and when breastfeeding based on animal studies. MC is also not recommended for humans with G6PD-deficiency.

## Data Availability

All data in the present study are available from previous publications.

https://pubmed.ncbi.nlm.nih.gov/

## List of abbreviations

AE: adverse events
AF: atrial fibrillation
ALT: alanine aminotransferase
AST: aspartate aminotransferase
BMI: body mass index
CAERS: centre adverse events reporting system
CI: confidence interval
CFSAN: Center for Food Safety and Applied Nutrition
EFSA: European food and safety authority
FDA: food and drug administration
FPG: fast plasma glucose
GABA: gamma-aminobutyric acid
GMP: good manufacturing practice
G6PD: glucose-6-phosphate dehydrogenase
g: gram
Hb: hemaglobin
HbA1c: glycosylated hemoglobin Type A1C,
IRR: incidence rate ratio
MC: Momordica charantia
mcIRBP-19: peptideB19 amino acids mcIRBP-19
mg: milligram
ml: milliliter
mmol/L: millimoles per litre
N: number
OAA: oral diabetic agent
PPG: postprandial glucose
ppm: parts per million
RCT: randomized controlled trial
TCM: traditional Chinese medicine
T2DM: type-2 diabetes mellitus
WHO: world health organization
wks: weeks
w/v: weight per volume
w/w: weight per weight

